# Improved characterization of diffusion in normal and cancerous prostate tissue through optimization of the restriction spectrum imaging signal model

**DOI:** 10.1101/2020.03.27.20042069

**Authors:** Christopher Charles Conlin, Christine H Feng, Ana E RodrÃ-guez-Soto, Roshan A Karunamuni, Joshua M Kuperman, Dominic Holland, Rebecca Rakow-Penner, Michael E Hahn, Tyler M Seibert, Anders M Dale

## Abstract

**Background:** Optimizing a restriction spectrum imaging (RSI) model for the prostate could lead to improved characterization of diffusion in the prostate and better discrimination of tumors.

**Purpose:** To determine optimal apparent diffusion coefficients (ADCs) for prostate RSI models and evaluate the number of tissue compartments required to best describe diffusion in prostate tissue.

**Study Type:** Retrospective.

**Population/Subjects:** Forty-six patients who underwent an extended MRI examination for suspected prostate cancer; 23 had prostate tumors and 23 had no detectable cancer.

**Field strength/Sequence:** 3T multi-shell diffusion weighted sequence.

**Assessment:** RSI models with 2-5 tissue compartments were fit to multi-shell DWI data from the prostate to determine optimal compartmental ADCs. Signal contributions from the different tissue compartments were computed using these ADCs and compared between normal tissues (peripheral zone, transition zone, seminal vesicles) and tumors.

**Statistical Tests:** The Bayesian Information Criterion (BIC) was used to evaluate the optimality of different RSI models. Model-fitting residual (as percent variance) was recorded to assess the models’ goodness-of-fit and whether it varied between anatomical regions of the prostate. Two-sample t-tests (α=0.05) were used to determine the statistical significance of any differences observed in compartmental signal-fraction between normal prostate tissue and tumors.

**Results:** The lowest BIC was observed from the 4-compartment model. Optimal ADCs for the 4 compartments were 5.2e-4, 1.9e-3, 3.0e-3, and ≫3.0e-3 mm^2^/s. Tumor tissue showed the largest reduction in fitting residual by increasing model order. Prostate tumors had a significantly (P≪0.05) greater proportion of signal from compartments 1 and 2 than normal tissue. Tumor conspicuity in compartment 1 increased substantially with model order.

**Data Conclusion:** Among the examined RSI models, the 4-compartment model best described the diffusion-signal characteristics of the prostate. Compartmental signal fractions revealed by such a model may improve discrimination between cancerous and benign prostate tissue.

## Introduction

Prostate cancer is the second most common malignancy in men worldwide, with over one million new diagnoses and 300,000 deaths annually (1, 2). While biopsy is the gold-standard technique for diagnosing prostate cancer, it is prone to sampling errors that can significantly impact risk stratification and treatment (3, 4). Multiparametric magnetic resonance imaging (MRI) has become an indispensable tool for improving diagnostic capabilities, aiding in the detection and characterization of prostate tumors, as well as providing image guidance for biopsy and focal intervention (5–8). A critical component of multiparametric MRI is diffusion-weighted MRI (DWI), which measures the diffusion properties of water at a microscopic level to assess the cytostructural makeup of tissue (9).

To identify cancerous lesions, conventional DWI analysis seeks to detect decreases in the apparent diffusion coefficient (ADC) of prostate tissue (5). However, the expected changes in ADC that accompany cancer are often confounded by edema or necrosis (10), and may not be detectable from ADC maps alone (11). It is particularly difficult to identify cancer in the transition zone of the prostate due to the common occurrence of benign prostatic hyperplasia, which exhibits DWI signal and ADC characteristics similar to that of tumors (12, 13).

Restriction spectrum imaging (RSI) is a more sophisticated DWI technique that employs a multi-shell diffusion acquisition and high b-values to account for cellular geometry and compartmentalization (10, 14, 15). RSI and other multi-shell DWI methods (16, 17) model the diffusion-weighted signal as a linear combination of exponential decays, with the individual decay-curves corresponding to different tissue compartments. The ADC value of each compartment is fixed, and variation in diffusion signal between voxels is therefore interpreted as variation in the proportion of each tissue compartment comprising the total diffusion signal. Fixing compartmental ADC values enables linearization of the DWI signal decay and rapid discrimination of the different tissue compartments within each voxel (14).

Meaningful assessment of prostate cancer with RSI requires that the number of signal-model compartments and their corresponding ADC values accurately characterize the diffusion properties of both normal and cancerous prostate tissue. Previous studies demonstrated improved discrimination between normal and cancerous prostate tissue using simple 2-compartment RSI models, with fixed ADC values corresponding to restricted and free diffusion of water (18–21), but stopped short of examining the optimality of such ADC values or comparing against higher-order models with additional tissue compartments. Optimizing the number of tissue compartments and associated ADCs of the RSI signal model could lead to improved characterization of prostate cancer and better discrimination of tumors in radiographically-complex regions like the transition zone.

In this study, we determined optimal ADC values for several RSI models of the prostate and assessed the number of tissue compartments required to best describe diffusion in both normal and cancerous prostate tissue. We then applied an optimized model to examine the diffusion profile of prostate cancer tumors and normal prostatic tissue, specifically the peripheral zone, transition zone, and seminal vesicles.

## Materials and Methods

This retrospective study was approved by the local institutional review board (IRB). A waiver of consent was obtained from the IRB to access patient MRI data and other clinical records. Forty-six patients who underwent an extended MRI examination for suspected prostate cancer were included. Standard-of-care evaluation determined that 23 of these patients (age: 64±9 years; PSA: 9.5±7.5 ng/mL) had cancerous lesions in the prostate, while the remaining 23 (age: 64±11 years; PSA: 12.2±23.8 ng/mL) had no detectable cancer.

Seventeen lesions were located (radiographically) in the peripheral zone of the prostate, 4 were located in the transition zone, and 2 extended into both the peripheral and transition zones. The PI-RADS scores for these lesions ranged from 3 to 5, with 13 lesions scored as a 5, 8 lesions scored as a 4, and 2 lesions scored as a 3. For pathologic examination of these radiographically identified lesions, all patients underwent systematic biopsy of the prostate. Nine patients additionally underwent targeted biopsies, and 12 patients ultimately underwent radical prostatectomy. Gleason Grade Groups reported for the lesions ranged from 1 to 5, with 4 lesions scored as a 5, 1 lesion scored as a 4, 7 lesions scored as a 3, 8 lesions scored as a 2, and 3 lesions scored as a 1. A more detailed summary of these clinical findings is presented in Table 1.

**Table 1:** Clinical findings from the 23 patients with prostate lesions included in this study. PZ: peripheral zone of the prostate, TZ: transition zone of the prostate, FIR: Favorable intermediate risk, UIR: Unfavorable intermediate risk, HR: High risk, VHR: Very high risk.

| Subject | Age | PI-RADS lesion score | Radiologic lesion location | PSA (ng/mL) | Gleason Grade Group | Pathology Specimen Type | Clinical Risk Group |
| --- | --- | --- | --- | --- | --- | --- | --- |
| 1 | 59 | 5 | Right TZ and PZ | 7.8 | 2 | Biopsy (Systematic) | FIR |
| 2 | 54 | 5 | Right PZ | 10.6 | 3 | Biopsy (Systematic),<br>Radical Prostatectomy | UIR |
| 3 | 71 | 5 | Right PZ and TZ | 4.0 | 5 | Biopsy (Systematic and Targeted),<br>Radical Prostatectomy | HR |
| 4 | 72 | 5 | Right PZ | 5.7 | 2 | Biopsy (Systematic) | UIR |
| 5 | 54 | 5 | Right PZ | 7.3 | 3 | Biopsy (Systematic),<br>Radical Prostatectomy | HR |
| 6 | 63 | 5 | Right PZ | 16.8 | 3 | Biopsy (Systematic),<br>Radical Prostatectomy | HR |
| 7 | 74 | 5 | Right PZ | 29.3 | 5 | Biopsy (Systematic) | HR |
| 8 | 53 | 4 | Left PZ | 8.0 | 1 | Biopsy (Systematic),<br>Radical Prostatectomy | FIR |
| 9 | 66 | 4 | Right PZ | 8.7 | 3 | Biopsy (Systematic and Targeted) | UIR |
| 10 | 67 | 5 | Left PZ | 4.6 | 2 | Biopsy (Systematic and Targeted) | FIR |
| 11 | 62 | 5 | Left PZ | 14.0 | 3 | Biopsy (Systematic),<br>Radical Prostatectomy | HR |
| 12 | 74 | 4 | Left PZ | 4.4 | 3 | Biopsy (Systematic and Targeted),<br>Radical Prostatectomy | UIR |
| 13 | 50 | 3 | Left PZ | 4.3 | 2 | Biopsy (Systematic),<br>Radical Prostatectomy | HR |
| 14 | 65 | 4 | Right TZ | 8.0 | 1 | Biopsy (Systematic) | LR |
| 15 | 81 | 5 | Anterior TZ | 8.5 | 2 | Biopsy (Systematic and Targeted) | FIR |
| 16 | 77 | 5 | Right PZ | 3.5 | 4 | Biopsy (Systematic),<br>Radical Prostatectomy | HR |
| 17 | 70 | 4 | Left PZ | 7.4 | 2 | Biopsy (Systematic and Targeted) | FIR |
| 18 | 58 | 4 | Left PZ | 7.4 | 2 | Biopsy (Systematic),<br>Radical Prostatectomy | UIR |
| 19 | 62 | 4 | Left PZ | 5.0 | 1 | Biopsy (Systematic) | LR |
| 20 | 68 | 4 | Right PZ | 5.9 | 5 | Biopsy (Systematic and Targeted),<br>Radical Prostatectomy | HR |
| 21 | 64 | 5 | Midline to Right TZ | 8.6 | 2 | Biopsy (Systematic and Targeted) | FIR |
| 22 | 51 | 5 | Diffuse PZ | 33.0 | 5 | Biopsy (Systematic) | VHR |
| 23 | 51 | 3 | Right TZ | 5.4 | 3 | Biopsy (Systematic),<br>Radical Prostatectomy | UIR |

### MRI data acquisition

MR imaging was performed on a 3T clinical scanner (Discovery MR750; GE Healthcare, Waukesha, WI) using a 32-channel phased-array body coil surrounding the pelvis. A multi-shell DWI volume was acquired for each patient that sampled 5 b-values (0, 200, 1000, 2000, and 3000 s/mm^2^) at 6 unique diffusion-encoding gradient directions (one average per direction, TR: 5000 ms, TE: 80 ms, resolution: 1.7×1.7 mm, matrix: 96×96 resampled to 128×128, slice thickness: 3 mm, fat saturation: on, acquisition time: 5 min). The b = 0 s/mm^2^ volumes were acquired using both forward and reverse phase encoding to allow for correction of B0-inhomogeneity distortions (22). For anatomical reference, a high resolution T2-weighted volume was acquired with scan-coverage identical to that of the multi-shell DWI volume (TR: 6225 ms, TE: 100 ms, resolution: 0.43×0.43 mm, matrix: 320×320 resampled to 512×512, slice thickness: 3 mm, fat saturation: off, acquisition time: 5 min).

### MRI data post-processing

All post-processing and analysis of MRI data was performed using custom programs written in MATLAB (The MathWorks, Inc; Natick, MA). The multi-shell DWI volumes were first corrected for distortions due to B0-inhomogeneity, gradient nonlinearity, and eddy currents (14, 22). Because noise in MR images can bias estimated DWI parameters (20, 23), signal intensity in the multi-shell DWI volumes was corrected to account for the presence of the noise floor. Briefly, the mean background signal intensity of each volume was estimated and then subtracted from the observed signal intensity of each voxel to obtain the corrected signal intensity (S_corr_). Isotropic diffusion in the prostate was assumed, so the 6 directional diffusion volumes at each b value were averaged together.

All tissue contouring was performed by a radiation oncologist and reviewed for accuracy by two board-certified radiologists using MIM software (MIM Software, Inc; Cleveland, OH). For patients without cancer, ROIs were defined to include the entire prostate and seminal vesicles. To allow for examination of signal from different anatomical regions, separate sub-ROIs were also defined over the peripheral and transition zones of the prostate, as well as the seminal vesicles. For patients with prostate cancer, ROIs were defined over the tumor in agreement with standard-of-care clinical assessment: lesion signal intensity on all available imaging modalities (principally T2-weighted and DWI) was considered alongside pathologic findings to determine the extent of the tumor ROI. All ROIs were examined by the two radiologists and, if necessary, the contours were adjusted to achieve consensus between readers. The finalized ROIs were then exported as binary masks in a MATLAB-readable format.

### RSI modeling

The RSI model is defined by the following formula:

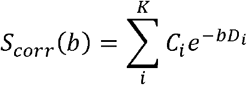

where S_corr_(b) denotes the noise-corrected DWI signal at a particular b value, K is the number of tissue compartments, C_i_ is a unit-less weighting factor describing the contribution of a particular compartment to the overall signal, and D_i_ is the compartmental ADC. By convention, the compartments are ordered from lowest to highest ADC, such that the first compartment describes the most restricted mode of diffusion. To determine optimal K and D_i_ values for the prostate, a global fitting of the model to the multi-shell DWI data from all voxels within all ROIs (normal+cancer tissue; >200,000 voxels) was performed, with K ranging from 2 to 5 (the maximum number of compartments was limited to the number of b-values employed during image acquisition). For each K value, model fitting was performed using a simplex search method (24) to minimize a cost function that quantifies the difference between observed and model-predicted signal values for all voxels simultaneously. To compute the cost function for a given set of D_i_ values, a nonnegative least-squares optimization (25) was necessary to estimate the corresponding C_i_ values and generate the model-predicted signal values. Minimizing this cost function returned optimal D_i_ values for each of the K compartments of the model. Unlike conventional voxel-wise fitting approaches that determine D_i_ for each voxel independently, the method outlined here determines globally optimal D_i_ values for the entire population of voxels across all patients (only the C_i_ weights are voxel-wise independent). This global fitting approach produces a highly overdetermined system that ensures stable estimates of D_i_ for all models examined in this study.

To determine how well the various multi-compartmental RSI models described the prostate diffusion data, the Bayesian Information Criterion (BIC; (26)) was computed for each model as follows:

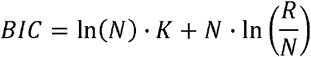

where N is the sample size (taken to be the number of patients in this study — 46), K is the number of tissue compartments in the model, and R is the model-fitting’s sum of squared residuals. A lower BIC indicates a better description of the data. The BIC value of a particular model is only meaningful in comparison to BIC values from other models under consideration, so in this study we reported only the relative BIC (ΔBIC) for each RSI model: Δ*BIC* = *BIC*_*i*_ − *BIC*_*min*_, where BIC_i_ is the BIC value of a particular model and BIC_min_ is the minimal BIC observed from any of the models.

Once optimal RSI models were determined, tissue signal-contribution (C_i_) maps were computed for each patient via nonnegative least-squares fitting of the model to the signal-vs-b-value curve from each voxel. Relative fitting residual (percent variance of the difference between model-predicted and measured signal) was recorded to assess the model’s goodness-of-fit and whether it varied between anatomical regions of the prostate. The distribution of signal-contribution among model compartments was compared between normal tissues (peripheral zone, transition zone, seminal vesicles) and tumors. Two-sample t-tests (α=0.05) were used to determine the statistical significance of any differences observed in compartmental signal-contribution between tissue types. Previous studies (10, 18–20) reported high tumor salience on the signal-contribution map of the compartment with lowest ADC (C_1_). Here we compared tumor conspicuity (ratio of mean signal in the tumor to mean signal in the surrounding parenchyma) between the C_1_ maps of the various models to assess the impact of RSI model order on tumor prominence.

## Results

Optimal ADC values for the different RSI models are listed in Table 2. Signal contribution (C_i_) maps that were computed from these optimized models are shown in Figure 1 for a subject with a primary tumor in the transition zone. The tumor appears as a bright region of the C_1_ map of each model. As model order increases, so too does the conspicuity of the tumor on the C_1_ map (Figure 2) as signal in the surrounding prostate decreases. The C_1_ map of the 2-compartment RSI model also shows signal contributions from throughout the prostate, while the C_2_ map is brightest in the peripheral zone of the prostate with additional signal contributions from the urethra and peripheral vessels. The 3-compartment model separates the signal contributions from normal (non-tumor) prostate and peripheral vessels into C_2_ and C_3_, respectively. In the 4-compartment model, normal prostate tissue is visible in C_2_ and C_3_, with peripheral vessels comprising C_4_. The 5-compartment model shows normal prostate tissue in C_2_, C_3_, and C_4_, with vasculature in C_5_.

**Table 2:** Optimal D_i_ values for RSI models of the prostate that describe 2 to 5 different tissue compartments. The relative Bayesian information criterion (ΔBIC) describes how well each model fits the data, with a lower ΔBIC indicating a better fit.

| Number of tissue compartments | Optimal $D_i$ for each compartment ($\text{mm}^2/\text{s}$) | | | | | $\Delta BIC$ |
| --- | --- | --- | --- | --- | --- | --- |
|  | 1 | 2 | 3 | 4 | 5 |  |
| 2 | 2.0e-3 | 5.2e-3 |  |  |  | 45.2 |
| 3 | 8.7e-4 | 2.6e-3 | 9.1e-3 |  |  | 11.8 |
| 4 | 5.2e-4 | 1.9e-3 | 3.0e-3 | $\gg 3.0\text{e-}3$ | | 0.0 |
| 5 | 0 | 1.3e-3 | 2.2e-3 | 3.1e-3 | $\gg 3.0\text{e-}3$ | 0.6 |

**Figure 1:**
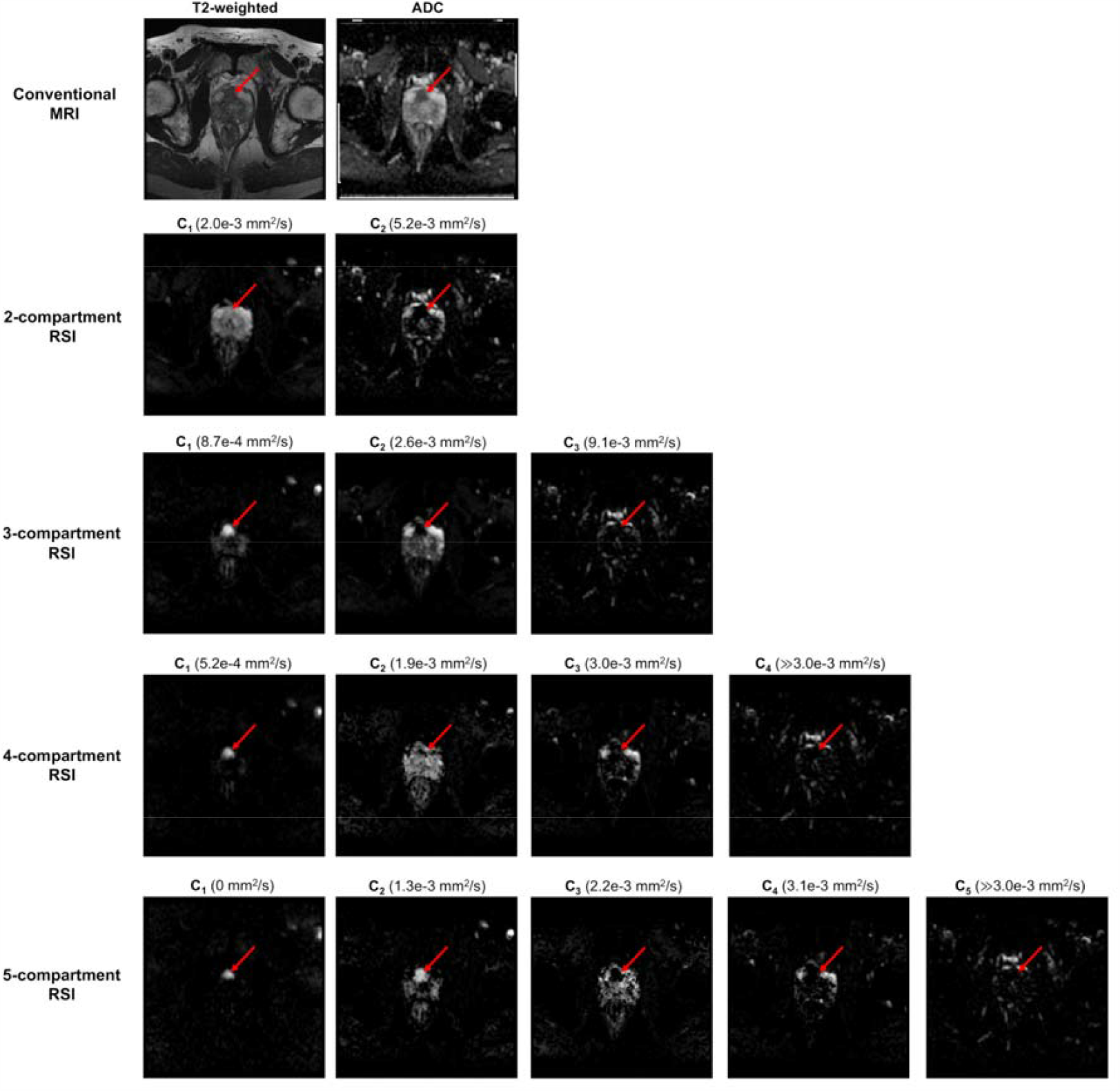
Axial images of the prostate from a patient with a primary tumor in the transition zone. A red arrow indicates the tumor in each image. Conventional T2-weighted and ADC images are shown in the top row, with RSI signal-contribution maps (C_i_) calculated from the optimized models in the following rows. The corresponding ADC of each compartment is listed in parentheses next to the compartment label. The 4- and 5-compartment RSI models show tumor signal contributions from multiple compartments, revealing diffusion-signal heterogeneity within the tumor that is not apparent in the conventional ADC image.

**Figure 2:**
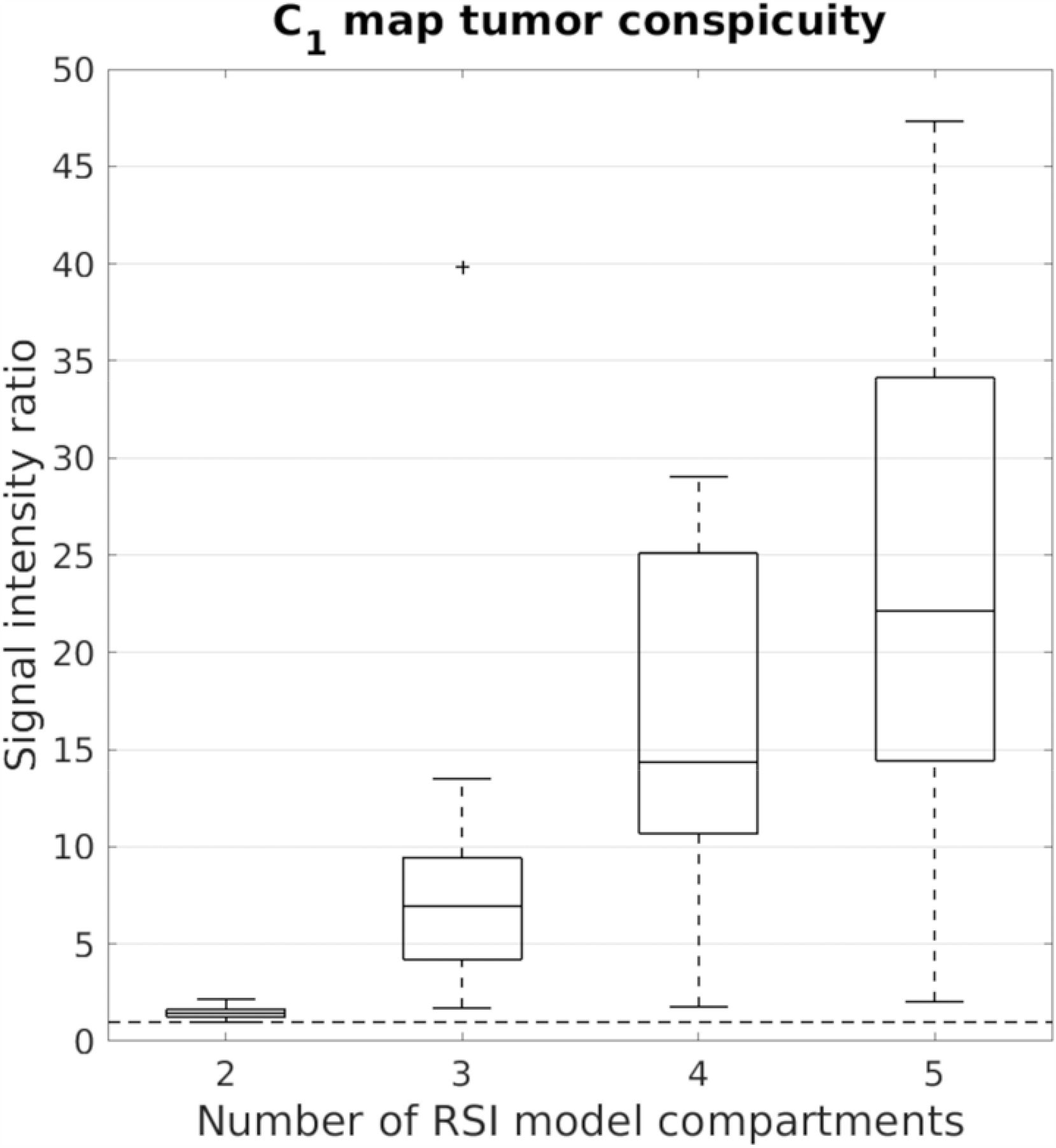
Tumor conspicuity on the C_1_ map of RSI models ranging from 2 to 5 tissue compartments. Tumor conspicuity increases substantially as model order increases. The dashed horizontal line marks the 1:1 tumor:parenchyma signal intensity ratio. A “+” above a box plot indicates an outlier.

The higher-order RSI models show tumor signal contributions from more than one compartment, revealing diffusion-signal heterogeneity within the tumor that is not apparent in the conventional ADC image. In the 4-compartment model, for instance, there is signal from the tumor on the C_2_ map as well as the C_1_ map. While C_1_ is hyperintense in the tumor alone, C_2_ is also bright throughout the transition zone. The 5-compartment model also shows tumor signal from C_2_ in addition to C_1_. Unlike the 4-compartment model, however, there is substantially less signal from the transition zone in C_2_ since the transition zone signal contribution is largely partitioned into C_3_ instead.

Signal contribution profiles of different tissues are quantified in Figure 3 for the various RSI models. Across all models, prostate tumors showed a significantly greater (P ≪ 0.05) proportion of signal from compartment C_1_ than was observed in normal tissue. With the 4- and 5-compartment RSI models, the fraction of signal from C_2_ was also significantly higher (P ≪ 0.05) in tumors compared to normal tissue, while that of C_3_ was significantly lower (P ≪ 0.05). Among normal tissues, the peripheral zone and transition zone had substantially different signal contribution profiles. With any model, the C_1_ signal fraction was significantly greater (P ≤ 0.01) in the transition zone than the peripheral zone. In the 2- and 3-compartment models, the C_2_ signal fraction was significantly larger in the peripheral zone (P ≤ 0.01). In the 4- and 5-compartment models, the C_2_ signal fraction was significantly higher in the transition zone (P ≪ 0.05). The 4-compartment model showed a significantly greater proportion of signal from C_3_ in the peripheral zone (P ≪ 0.05). The signal proportions computed from the 5-compartment model differed significantly between the peripheral and transition zones in compartments C_3_ (P = 0.01) and C_4_ (P ≪ 0.05). In the final compartments of the 4-, and 5-compartment RSI models (which have the highest D_i_ values and correspond to the vessel images in Figure 1) the signal fraction from the transition zone was significantly larger than from the peripheral zone (P ≤ 0.02). None of the compartmental signal fractions were significantly different between the peripheral zone and seminal vesicles (P > 0.1 for all comparisons of all models).

**Figure 3:**
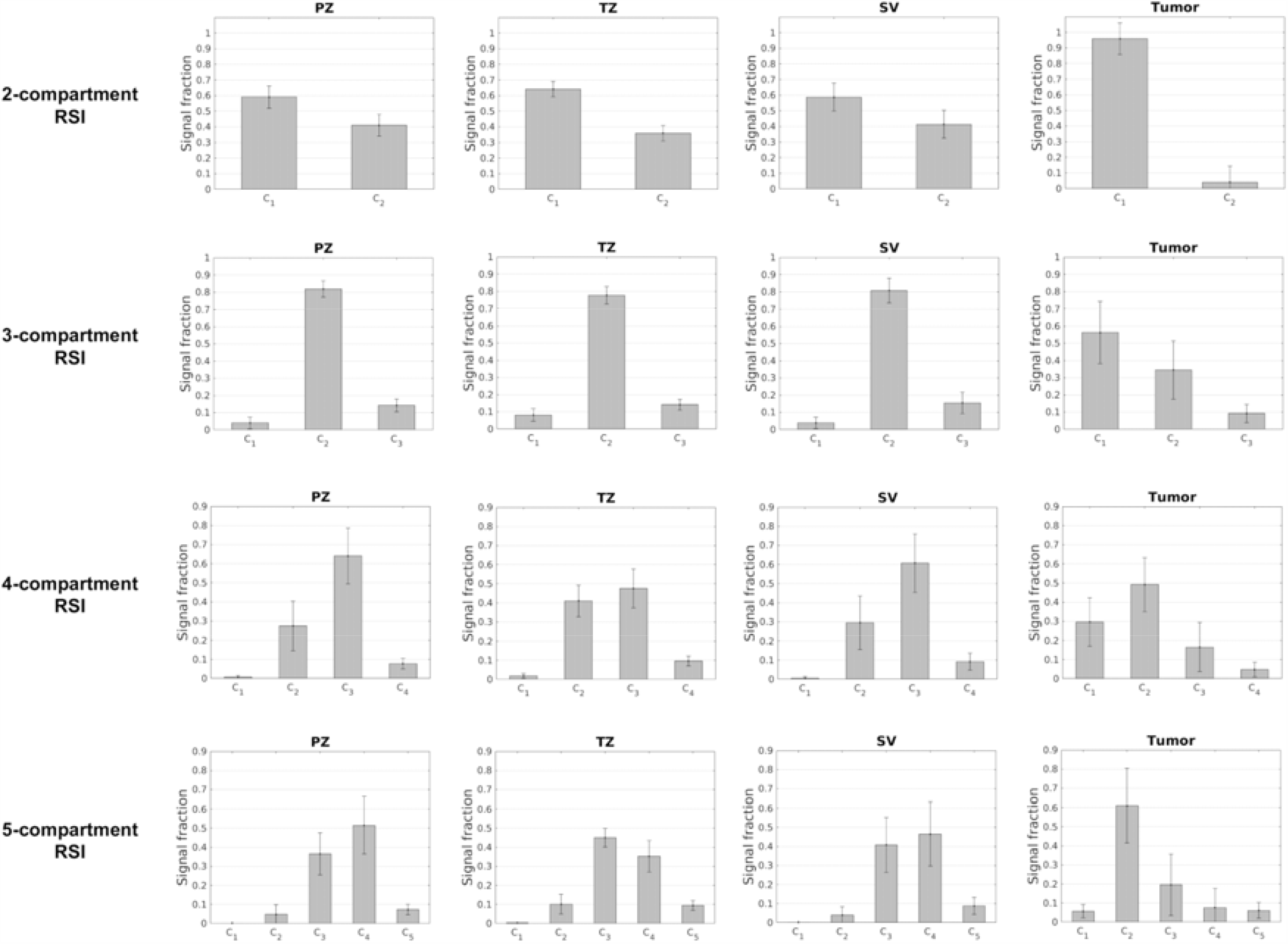
RSI signal-contribution profiles of different tissues, with models ranging from 2 to 5 compartments. PZ: peripheral zone of the prostate, TZ: transition zone of the prostate, SV: seminal vesicles.

The relative BIC (ΔBIC) for each RSI model is listed in Table 2. ΔBIC values were substantially lower for the 4- and 5-compartment models than for the 2- and 3-compartment models. The lowest BIC was observed from the 4-compartment model, suggesting that it provides the optimal characterization (among the models examined in this study) of diffusion properties throughout the entire prostate and seminal vesicles. Figure 4 compares the fitting residuals by tissue type between the different models and demonstrates the improved fit of the model to the data (i.e., reduced fitting residual) with increasing model order. As model order was increased from 2- to 5-compartments, the fitting residual (in percent variance) decreased by 0.15 across all voxels, 0.11 in the peripheral zone, 0.11 in the transition zone, 0.14 in the seminal vesicles, and 4.57 in tumors. Tumor tissue showed the largest reduction in fitting residual by increasing model order.

**Figure 4:**
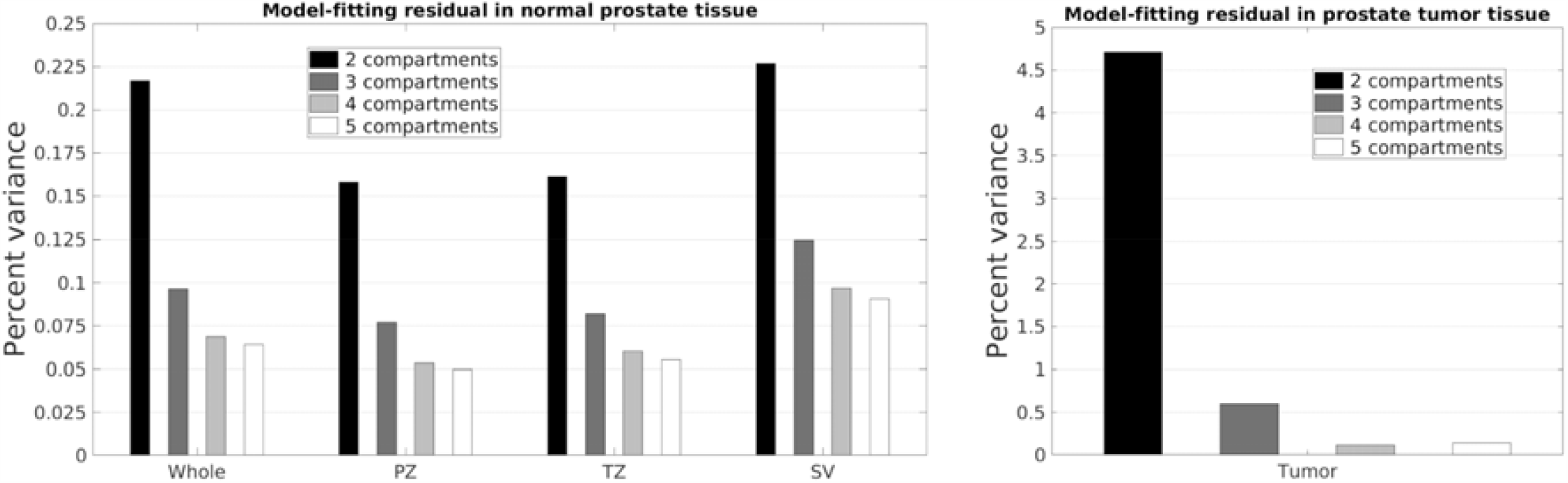
RSI model-fitting residual by tissue type. Residuals in normal prostate tissue are shown on the left, while residuals in tumor tissue are shown on the right. Whole: whole prostate plus seminal vesicles, PZ: peripheral zone of the prostate, TZ: transition zone of the prostate, SV: seminal vesicles.

## Discussion

In this study, we evaluated different RSI models to determine an optimal characterization of diffusion properties in both normal and cancerous prostate tissue. The diffusion properties (D_i_) of each model were determined through simultaneous, global fitting to diffusion-signal data from 46 clinically-representative patients (over 200,000 voxels). Unlike more conventional DWI analysis that examines only the averaged signal within pre-identified lesion ROIs (27–29), our approach considers the diffusion properties of every tissue voxel, whether it is cancerous or not, throughout the entire prostate and seminal vesicles. Other multi-shell DWI methods have examined diffusion at the voxel level (16, 17), but the RSI approach employing fixed, globally-optimal D_i_ values enables more stable estimates than can be achieved with fully-independent model fitting at each voxel. Enhanced estimation stability is particularly important when fitting higher-order multicompartmental models as was done in this study.

Among the signal models examined, which ranged from 2 to 5 tissue compartments, the 4- and 5-compartment models showed substantially lower ΔBIC values compared to the 2- or 3-compartment models. Such reduced ΔBIC suggests that these higher-order models provide a better description of diffusion signal from the prostate. While our results show that RSI models with 2 or 3 compartments may be suitable for gross detection of prostate tumors, they lack sufficient resolution to distinguish between the different diffusional modes within the tumor that were revealed using higher-order models (Figure 1). Furthermore, the improvement in tumor conspicuity observed with increasing model order (Figure 2) may be a clinically-relevant benefit that could potentially outweigh the increase in scan time required to obtain data at the additional b-values needed to implement these models. Given the b-values of the data that were available for this retrospective study, we were unable to examine RSI models with more than 5 compartments. However, the optimal ΔBIC observed from the 4-compartment model suggests that the addition of more compartments will not provide significant additional benefit.

Looking at the optimal D_i_ values for the 5-compartment model, we can begin to discern the different modes of diffusion that contribute to DWI signal in the prostate. The optimal D value of compartment C_1_, 0 mm^2^/s, suggests that it reflects signal contributions from highly restricted cellular structures that have no detectible diffusion over the effective diffusion times used in this study (14). The D of compartment C_2_, 8.9e-4 mm^2^/s, is consistent with the characterization of restricted diffusion from previous studies (14, 18–20), ostensibly representing intracellular diffusion. Compartment C_3_ accounts for hindered diffusion through the extracellular extravascular space (14, 30), having an optimal D of 1.7e-3 mm^2^/s. Compartment C_4_, with an optimal D of 2.7e-3 mm^2^/s, reflects the free diffusion of water (14, 31). Finally, the fifth compartment C_5_ considers rapid pseudo-diffusion (IVIM effects (32)) with an optimal D much greater than that of free diffusion (≫3.0e-3 mm^2^/s).

The distribution of these modes of diffusion in both normal and cancerous prostate tissue was revealed (Figure 3) by fitting the RSI models to multi-shell prostate diffusion data, and may provide insight into the cytostructural changes that accompany prostate cancer development. Of particular clinical interest is the increased cellularity of tumors relative to normal tissue, which is an important prognostic indicator for prostate cancer (21, 33). Previous RSI studies have attempted to measure restricted diffusion within cells to assess tumor cellularity (18–21), and have reported a correlation between RSI signal contribution and histological Gleason grades. However, the signal models in these studies were limited to only two tissue compartments with fixed D_i_ values reflecting restricted and free diffusion. While the signal fractions presented in Figure 3 do indeed suggest that a 2-compartment model is sufficient to differentiate between cancerous and normal prostate tissue, it is clear that the granularity of analysis is limited with only two compartments since signal from the other modes of diffusion are forced into either the “restricted” or “free” compartments of the model.

Interestingly, the “restricted” compartment in these previous studies was assigned a D_i_ value on the order of 10^−4^ mm^2^/s, while the ADC of *truly* restricted diffusion within cells should be essentially zero over the relatively long effective diffusion times achievable on clinical MRI hardware (14, 34). It is likely that this compartment did not actually represent pure intracellular diffusion, but rather a mixture of intracellular diffusion and highly hindered or tortuous extracellular diffusion (34). The 5-compartment RSI model enables differentiation between these two modes of diffusion, which might help to discriminate between cancerous and benign tissue in radiographically-complex regions like the transition zone. While the ΔBIC values reported here suggest that the 4-compartment model, which lacks a compartment for purely restricted diffusion, is slightly more optimal than the 5-compartment model, this may be due to the relatively small number of transition zone tumors in our dataset. Subsequent studies will evaluate the efficacy of these higher-order models to describe cancer in the transition zone specifically.

This study has a few limitations. First, the generalizability of our findings may be limited since data was obtained from a relatively small number of subjects at a single institution. However, the subjects that were included are largely representative of the broader clinical population, and there was sufficient statistical power for the reported hypothesis testing. Another limitation is that this study evaluated the different models only in terms of their ability to fit diffusion data (i.e., using BIC and fitting residual), and not their actual sensitivity and specificity for prostate tumors. While the higher-order models showed a better fit to the diffusion data, it is not clear whether they would outperform simpler models in terms of classifying tissue as cancerous or benign. Nevertheless, the results of this current study are still valuable for developing a comprehensive model of diffusion in the prostate. Subsequent studies will evaluate RSI models for prostate cancer screening specifically.

The RSI model-optimization procedure outlined here can be readily applied to tissues other than the prostate. As a straightforward extension of this study, we optimized RSI models for all tissues included in the original imaging volume, not just the prostate and seminal vesicles (see Supplementary Material online). We hypothesize that such models will better characterize the diffusion of both normal and malignant tissue throughout the body, potentially leading to improved identification of cancer in tissues beyond just the prostate. Future work will focus on leveraging these models to develop automated cancer screening methods.

## Data Availability

Contact the corresponding author with any inquiries regarding the data presented in this manuscript.

## Notes

### Competing Interest Statement

Anders M Dale reports that he was a Founder of and holds equity in CorTechs Labs, Inc., and serves on its Scientific Advisory Board. He is a member of the Scientific Advisory Board of Human Longevity, Inc. He receives funding through research grants from GE Healthcare to UC San Diego. The terms of these arrangements have been reviewed by and approved by UC San Diego in accordance with its conflict of interest policies.

### Funding Statement

This work was supported by funding from the following sources:
USAMR DoD W81XWH-17-1-0618
NIH K08 NIBIB EB026503
Prostate Cancer Foundation
UC San Diego Center for Precision Radiation Medicine

